# The Right Tool for the Task: Body-Weight Supported Treadmill or Total Body Recumbent Stepper for Mobility-Adapted Cardiopulmonary Exercise Testing in Multiple Sclerosis Patients with Disability

**DOI:** 10.1101/2024.12.11.24318556

**Authors:** Saman Hadjizadeh Anvar, Liam P Kelly, Caitlin Newell, Lynsey Alcock, Michelle Ploughman

**Author notes:** Corresponding Author: Michelle Ploughman, Recovery and Performance Laboratory, Faculty of Medicine, Memorial University of Newfoundland, Rm 400, L. A. Miller Centre, 100 Forest Rd, St. John’s, NL, A1A 1E5, Canada.

## Abstract

**Objective:** Cardiopulmonary Exercise Testing (CPET) is challenging among persons with mobility disability. We sought the optimal adapted device to achieve a maximal CPET.

**Design:** Randomized crossover trial, within-subjects, repeated measures design

**Setting:** Primary Care and Referral Center

**Participants:** Clinic-referred persons with multiple sclerosis (PwMS) (n=10) with three-month stability, no exercise obstruction, MoCa>24, ability to walk with or without assistance, and sex- and age-matched (±3 years) Controls (n=7) recruited by convenience sampling

**Interventions:** CPET on body weight-supported treadmill (BWST) and total body recumbent stepper (TBRS)

**Main Outcome Measures:** Standard aerobic metrics (V̇O_2max_, % normative values for V̇O_2max_ [%V̇O_2max_], heart rate maximum [HR_max_], age-predicted HR_max_, and Respiratory Exchange Ratio)

**Results:** PwMS achieved similar V̇O_2max_ (mL·min^-1^·kg^-1^) on the TBRS and BWST (26.53±8.7 vs. 24.24±7.8) while Controls obtained higher values on BWST than TBRS (40.27±7.6 vs. 34.32±7.1, *p*<0.001). PwMS more consistently achieved criteria for maximum CPET using TBRS. During the preliminary investigation of the MS subgroup with a higher mobility disability, CPET using BWST exaggerated already low CPET metrics.

**Conclusions:** Although Controls achieved higher CPET values on BWST, V̇O_2max_ between devices were similar among PwMS. Only when using BWST, PwMS V̇O_2max_ and %V̇O_2max_ were lower than Controls, likely because of leg fatigue and weakness. Using TBRS permits persons with mobility disability to achieve more criteria for a maximum CPET. Our results suggest that CPET using BWST, being reliant on the lower body, likely disadvantages PwMS, especially those with mobility disability.

## INTRODUCTION

Multiple sclerosis (MS) is the most prevalent neurological disorder among young and middle-aged people [<u>1</u>]. Adopting an inactive lifestyle [<u>2</u>] results in inactivity-related health problems among persons with MS (PwMS), such as osteoporosis, cardiovascular disease, fatigue, and depression [<u>3</u>]. Aerobic capacity is a crucial health [<u>4</u>] and performance marker [<u>5</u>] [<u>6</u>], and PwMS exhibit lower values of maximal aerobic power when compared to age and sex-matched healthy adults [<u>7</u>]. Importantly, lower risk of cardiovascular disease [<u>6</u>], better walking performance [<u>8</u>], faster cognitive processing speed [<u>9</u>], and enhanced neuroprotection and brain health [<u>10</u>], have all been associated with greater aerobic capacity in MS [<u>6</u>]. Testing aerobic fitness accurately, as a metric to calculate personalized exercise intensity and as a study outcome measure, is recommended [<u>6</u>].

The gold standard assessment of aerobic fitness measures the maximum rate of oxygen consumption (V̇O_2max_), in relation to the rate of carbon dioxide (V̇CO_2_) produced during a progressive maximal cardiopulmonary exercise test (CPET). CPET is feasible and safe in PwMS [<u>11-13</u>]. While several different exercise testing modalities (e.g., treadmill, bicycle ergometer, recumbent stepper) have been used to assess aerobic capacity, there is no evidence-based consensus regarding the best instrument that should preferentially be used in people having mobility disability [<u>13</u>].

Weakness and fatigue of the limbs can limit the ability to reach maximum aerobic capacity during a CPET [<u>14</u>, <u>15</u>]. Additionally, leg weakness and fear of falling could impede maximal effort, even when using treadmills with overhead harness support [<u>16</u>]. Although yet to be determined, modalities that permit the user to exercise in a seated position, such as total body recumbent steppers (TBRS), may permit CPET metrics that represent true cardiopulmonary capacity, less influenced by leg impairments [<u>17</u>]. Obtaining stable and representative CPET values would be especially useful in longitudinal studies, as there may be an emergence of MS-related relapses or accumulation of disability within an individual.

Investigators have tested several adapted and non-adapted methods for CPET among PwMS, including bicycle ergometers, arm ergometers, and TBRS [<u>11</u>, <u>18</u>]. For instance, Pilutti and colleagues compared arm ergometers to TBRS, and reported higher peak aerobic capacity obtained with TBRS, especially among people with low levels of disability and higher fitness levels [<u>19</u>]. Although useful in determining optimal CPET methodology, the authors did not provide comprehensive reporting of criteria necessary for V̇O_2max_ [<u>19</u>, <u>20</u>], so whether maximum CPET was achieved cannot be assured. The researchers recommended the use of a TBRS over the arm ergometer for CPET in PwMS. Similarly, evidence from our laboratory, testing two common adapted CPET methods (TBRS and Body Weight Supported Treadmill [BWST]) suggested less fatigue of the soleus muscle when the workload was distributed between four limbs (using a TBRS) rather than two (using a BWST) [<u>21</u>].

We aimed to compare CPET metrics using two adapted modalities (BWST and TBRS) among PwMS and matched controls. Metabolic parameters (V̇O_2max_, % normative values for V̇O_2max_ [% V̇O_2max_], heart rate maximum [HR_max_], and age-predicted HR_max_) were obtained along with the ability of participants to meet criteria for achieving a maximum CPET[<u>22</u>]. We also aimed to determine, in a preliminary way, whether the level of disability influenced the capacity to achieve the indicators of V̇O_2max_. We hypothesized that: a) Controls would achieve similar, if not higher, V̇O_2max_ on the BWST and TBRS; b) PwMS would achieve higher values and be more likely to meet V̇O_2max_ criteria on TBRS, especially for those having greater mobility disability.

## METHODS

### Participants

Based on previous studies [<u>23</u>] [<u>24</u>], ten PwMS were recruited as part of a study to examine the effects of BWST or TBRS on lower limb fatigue [<u>21</u>]. PwMS were recruited from an MS Neurology clinic and outpatient rehabilitation services. To be included, we confirmed: a) MS diagnosis using McDonald criteria [<u>25</u>];b) no relapses/stable during the previous three months; c) a negative Physical Activity Readiness Questionnaire (PAR-Q) screening [<u>26</u>]; d) no musculoskeletal obstruction to exercise; and e) scoring Montreal Cognitive Assessment (MoCA) > 24 [<u>27</u>]; f) ability to walk at least 10m with or without assistance. Sex- and age-matched (±3 years) Controls were recruited by convenience sampling. Competitive athletes were excluded [<u>28</u>]. After obtaining written informed consent, sex, age, height, weight, Expanded Disability Status Scale (EDSS), type of MS (relapsing-remitting [RRMS], secondary progressive [SPMS], or primary progressive [PPMS]), medications, and co-morbid conditions were recorded. The Health Research Ethics Board (HREB) of Memorial University of Newfoundland gave ethical approval for this study (Approval Ref. #: 14.102).

### Experimental Design

In a randomized crossover trial, within-subjects, repeated measures design, participants performed the CPET on the BWST and TBRS, 7-10 days apart at the same time of day with five minutes each of warm-up and cool-down. Participants were required to avoid food, caffeine, and intense exercise for at least two, six, and 12 hours, respectively, before the tests [<u>20</u>].

### Interventions

#### Total Body Recumbent Stepper (TBRS)

Using NuSTEP T4r Recumbent Stepper (NuStep Inc, Ann Arbor, MI) in a seated position [<u>12</u>, <u>29</u>], participants were required to maintain a speed of 80 strides per minute. The resistance (1-10, beginning at level 3 = 25 watts[<u>30</u>]) was increased by one unit (= 15 watts) every two minutes [<u>30</u>]. If the participant did not reach exhaustion at the highest resistance level (Level 10), the speed (strides per minute) was increased by ten every two minutes until volitional exhaustion terminated the test.

#### Body Weight Support Treadmill (BWST)

A rehabilitation treadmill (Sport Art T625M/T52 MD-Rehabilitation Commercial Treadmill, USA) was used with an overhead support harness at 10% of body weight. CPET started at a self-selected speed for two minutes with a %0 treadmill grade. While keeping a constant speed, the grade was increased by 2.5% every two minutes until the grade reached 10%. Reaching grade 10, the speed increased by 0.05 m/s every two minutes until volitional exhaustion.

After calibration, indirect calorimetry (Moxus Metabolic Systems, AEI Technologies, Inc., Pittsburgh, Pennsylvania, USA) was used to measure the rate of oxygen consumption (V̇O2), carbon dioxide production (V̇CO_2_), and HR (HR: Polar V800, Polar Electro Oy, Professorintie 5, FI-90440, Kempele, Finland). We recorded criteria for the termination of the CPET: (i) RPE > 7/10; (ii) no HR or V̇O_2_ increase despite increases in workload; (iii) inability to maintain the required workload or speed. Achievement of maximal oxygen consumption was assessed based on the attainment of 2 or more of the following criteria: (a) plateau in V̇O_2_ (≤150 ml/min^-1^), (b) RER ≥ 1.1; and/or (c) HR_max_ ± 10 beats per minute (bpm) of age-predicted HR_max_ based on the following equation: 206.9 – (0.67 * age) or 164 – (0.7 * age) if β-blockers prescribed [<u>29</u>, <u>31</u>]. Relative V̇O_2max_ was calculated with the highest absolute V̇O_2_ divided by the body weight and reported as ml.min^-1^.kg^-1^. V̇O_2max_ was also converted to normative values (% V̇O_2max_) based on the American College of Sports Medicine (ACSM) Guidelines for Exercise Testing and Prescription [<u>22</u>]. The time to test completion was also recorded in minutes.

### Statistical Analyses

Data were analyzed using SPSS version 27 software. Normality (Shapiro-Wilk test) and sphericity (Mauchly test) were confirmed. Greenhouse-Geisser epsilon was reported in the case of sphericity violation. Repeated measures ANOVA tests were used to detect any potential differences between MS and Controls in any of the dependent variables (V̇O_2max_, % V̇O_2max_, HR_max_, age-predicted HR_max_) on both modalities (TBRS, BWST), and the interaction effect (modality [TBRS vs. BWST] * Group [MS vs. Control]). One-way ANOVA tests were also used to compare disability level groups (> 2 and ≤ 2 EDSS) with Control on BWST and TBRS. Chi-square test was used to compare the number of exercise criteria achievement on TBRS and BWST (MS [EDSS > 2 & EDSS ≤ 2] vs. Control). Each participant’s achievement of fitness criteria on each modality was recorded as “Yes” or “No.” A significance level of α = 0.05 was chosen to assess the statistical significance of all testing variables. Effect sizes were reported as partial eta-squared (µ^2^), with 0.02, 0.13, and 0.26 considered small, medium, and large effects, respectively [<u>32</u>].

## RESULTS

### Participants

The median (IQR) for the PwMS EDSS score was 2.25 (6), ranging from 0 to 6.0, with half scoring 2.5 (minimal disability) or greater. On average, the PwMS ranged in age from 27-63 years. Groups did not differ in age (*p* = 0.9), or BMI (*p* = 0.1). Individual-level and summarized demographic and metabolic data are provided in tables 1 and 2, respectively. Controls did not differ in time to test completion between BWST and TBRS (*p* = 0.5). However, PwMS exercised about 9 minutes longer on BWST compared to TBRS (*p* < 0.001).

**Table 1.** Participant Demographics.

|  | ID | Sex | Age Range | BMI (kg/m<sup>2</sup>) | EDSS | MS Type | MS medication |
| --- | --- | --- | --- | --- | --- | --- | --- |
| MS Patients | 1 | Female | 51-55 | 35.4 | 6.0 | PPMS | MFS<sup>b</sup> |
|  | 2 | Male | 51-55 | 28.6 | 3.5 | RRMS | DMD<sup>a</sup> |
|  | 3 | Female | 56-60 | 21.9 | 3.0 | RRMS | DMD<sup>a</sup> |
|  | 4 | Female | 56-60 | 27.3 | 3.0 | RRMS | DMD<sup>a</sup> |
|  | 5 | Female | 46-50 | 31.7 | 2.5 | RRMS | DMD<sup>a</sup> |
|  | 6 | Female | 36-40 | 20.5 | 2.0 | RRMS | DMD<sup>a</sup> |
|  | 7 | Female | 26-30 | 19.2 | 1.0 | RRMS | DMD<sup>a</sup> |
|  | 8 | Female | 61-65 | 23.5 | 1.0 | RRMS | DMD<sup>a</sup> |
|  | 9 | Female | 46-50 | 24.5 | 0 | RRMS | None |
|  | 10 | Female | 41-45 | 34.7 | 0 | RRMS | None |
| **Mean ± SD:** |  |  | **49.2 ± 10.8** | **26.7 ± 5.7** | **2.2 ± 1.8** | N/A | N/A |
| Healthy Controls | 1 | Female | 51-55 | 19.6 | None | None | None |
|  | 2 | Female | 56-60 | 21.9 | None | None | None |
|  | 3 | Female | 41-45 | 22.7 | None | None | None |
|  | 4 | Female | 46-50 | 24.4 | None | None | None |
|  | 5 | Male | 51-55 | 25.2 | None | None | None |
|  | 6 | Female | 26-30 | 24.6 | None | None | None |
|  | 7 | Female | 61-65 | 24.8 | None | None | None |
| **Mean ± SD:** |  |  | **50.3 ± 11.3** | **23.3 ± 2.03** | N/A | N/A | N/A |

EDSS, Expanded Disability Status Scale; RRMS, Relapsing Remitting MS; PPMS, Primary Progressive MS; <sup>a</sup>, disease-modifying drug; <sup>b</sup>, medication for spasticity.

**Table 2.**
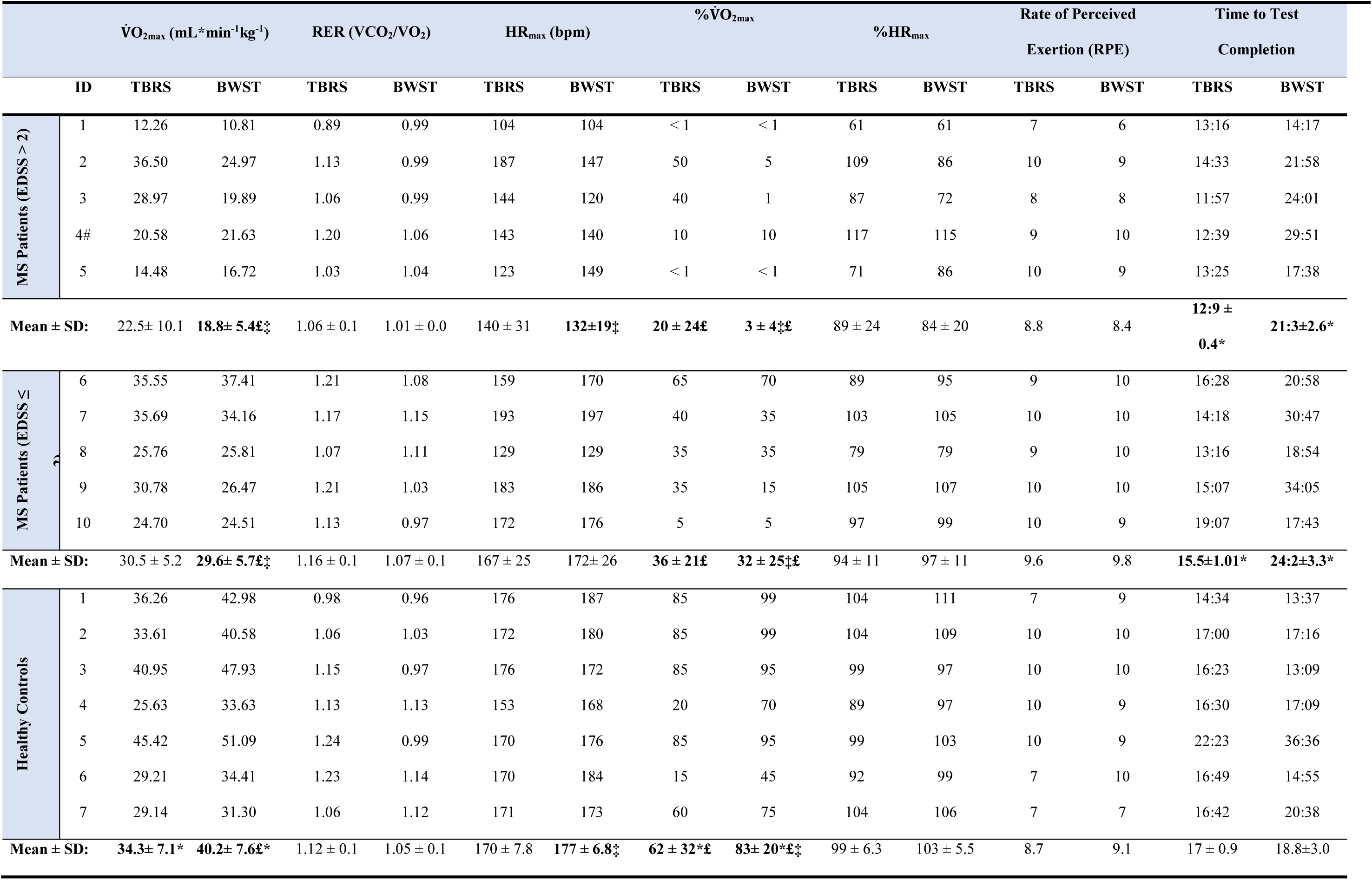
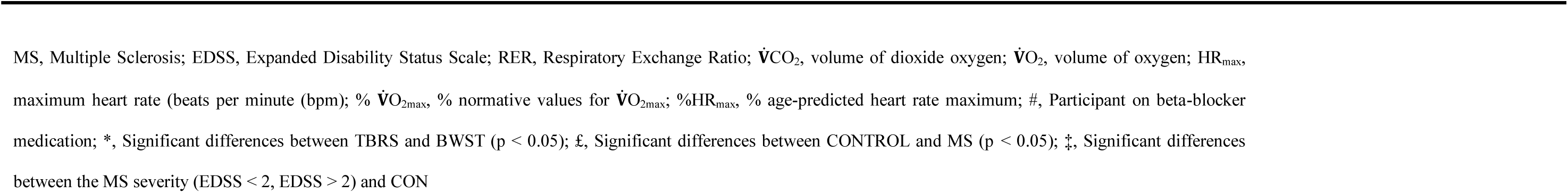
Individual Metabolic Parameters Obtained.

### Metabolic Parameters

#### V̇O_2max_

As expected, the Control Group’s V̇O_2max_ was significantly higher than PwMS overall, as a main effect was observed for the group (MS vs. Control) with a large effect size (F _(1, 15)_ = 10.0, *p* = 0.006, µ^2^ = 0.4; table 3 and figure 1). Although the PwMS achieved similar V̇O_2max_ using BWST and TBRS, the Control group’s V̇O_2max_ was higher on the BWST than when using the TBRS (Interaction effect, F _(1,15)_ = 19.3, *p* < 0.001, µ^2^ = 0.5) (figure 1). PwMS had lower V̇O_2max_ than Controls on the BWST (24.2±7.8 vs. 40.2±7.6 mL·min^-1^·kg^-1^) but not on the TBRS (26.5±8.7 vs. 34.3±7.1 mL·min^-1^·kg^-1^). A preliminary analysis suggested differences between the Controls and MS disability subgroups were more apparent when using BWST, likely because the Controls obtained higher V̇O_2max_ on BWST. (table 3; figure 2). After V̇O_2max_ values were transformed into % normative values for age and sex (table 3, figures 3 & 4), similar results were observed. Controls achieved higher values than PwMS (F_(1,15)_ = 19.2, µ^2^ = 0.56, *p* < 0.001). Controls obtained 83% normative values using BWST, significantly higher than TBRS (62%; F _(1,15)_ = 13.9, *p* = 0.002, µ^2^ = 0.48). PwMS performed similarly on the BWST (18%) versus the TBRS (28%), both significantly below the BWST and TBRS obtained from Controls (figure 3). When examining the MS disability subgroups, Controls were higher than both EDSS ≤ 2 (*p* = 0.001) and EDSS > 2 groups (*p* < 0.001) on BWST but not using TBRS (figure 4).

**Figure 1.**
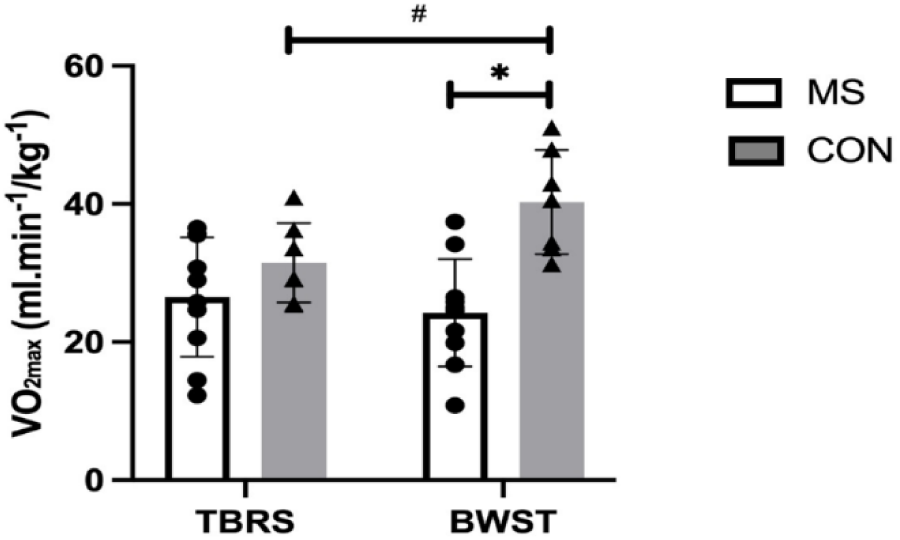
*V̇*O_2max_ in Multiple Sclerosis and Control (mean ± SD) when both groups performed CPET on BWST and TBRS. The Control group achieved a significantly higher *V̇*O_2max_ on the BWST (#; *p* < 0.001); Multiple Sclerosis participants had a significantly lower *V̇*O_2max_ than the Control on BWST (*; *p* < 0.001), but not on the TBRS.

**Figure 2.**
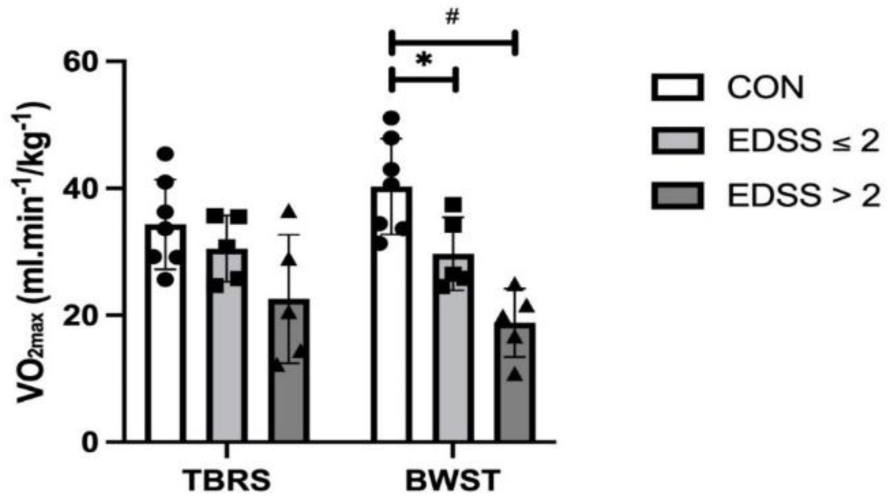
*V̇*O_2max_ differences (mean ± SD) between the two Multiple Sclerosis-severity participants (EDSS ≤ 2, EDSS ˃ 2) and Control. Participants with higher Multiple Sclerosis severity (EDSS > 2) were significantly lower than the Control, only on BWST (#, *p <* 0.001*, 95% C.I. = 11.1, 31.7*). Multiple Sclerosis participants with lower severity (EDSS ≤ 2) were also lower than the Control only on BWST (*, *p =* 0.04*, 95% C.I. = 0.27, 20.9*).

**Figure 3.**
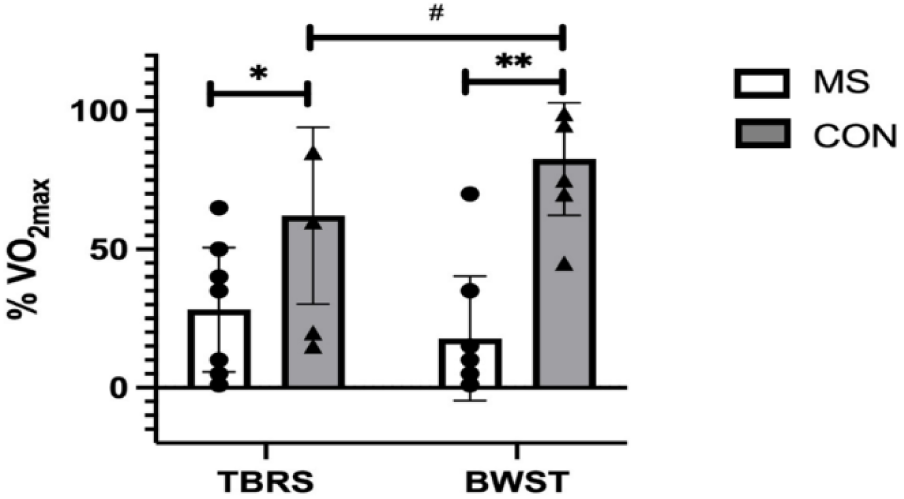
%*V̇*O_2max_ (% normative values) in Multiple Sclerosis and Control; the Control group was significantly higher on BWST than TBRS (#, p = 0.002). MS was lower than the Control on both BWST (**, *p* < 0.001) and TBRS (*, *p* = 0.02) (F _(1, 15)_ = 13.9, *p* = 0.002, µ^2^ = 0.48).

**Figure 4.**
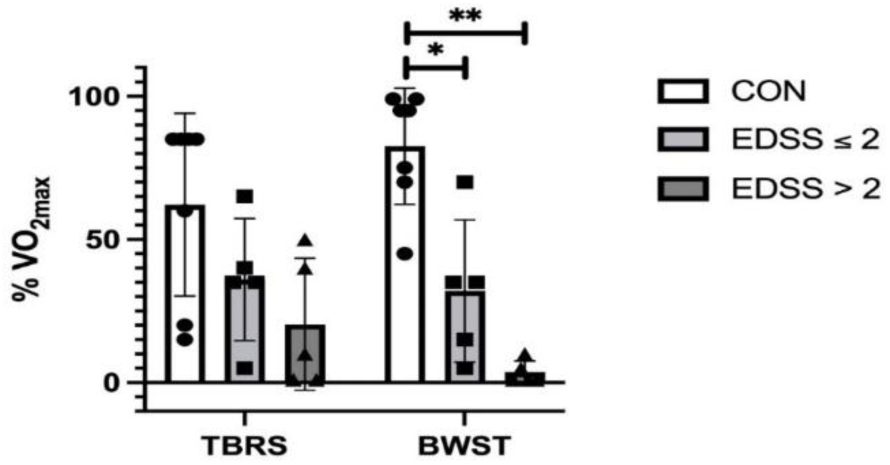
*V̇*O_2max_ (% normative values) differences between the groups of Multiple Sclerosis participants (EDSS ≤ 2; EDSS >2) and the Control. The Multiple Sclerosis participants with higher Multiple Sclerosis severity (EDSS >2) had significantly lower values than the Control, only on BWST (**, *p* < 0.001, 95% C.I. = 49.2, 109.5). Also, Multiple Sclerosis participants with lower Multiple Sclerosis severity (EDSS ≤ 2) had significantly lower values than the Control only on BWST (*, *p* = 0.001, 95% C.I. = 20.4, 80.7).

**Table 3.** Summary of Metabolic Parameters and Criteria for Achieving V̇O_2max_ on TBRS and BWST.

|  | MS Patients | MS Patients | MS Patients | Healthy Controls |
| --- | --- | --- | --- | --- |
|  | All | EDSS > 2 | EDSS ≤ 2 |  |
| Metabolic Parameters, TBRS |  |  |  |  |
| $\dot{V}O_{2max}$ (mL·min<sup>-1</sup>·kg<sup>-1</sup>) | 26.53 ± 8.7 | 22.56 ± 10.1 | 30.50 ± 5.2 | **34.32 ± 7.1\*** |
| RER (VCO<sub>2</sub>/VO<sub>2</sub>) | 1.11 ± 0.1 | 1.06 ± 0.1 | 1.16 ± 0.1 | 1.12 ± 0.1 |
| HR<sub>max</sub> (bpm) | 154 ± 30 | 140 ± 31 | 167 ± 25 | 170 ± 7.8 |
| % $\dot{V}O_{2max}$ | **28 ± 23£** | 20 ± 24 | 36 ± 21 | **62 ± 32\*£** |
| %HR<sub>max</sub> | 91.69 ± 17.8 | 88.95 ± 24.1 | 94.43 ± 10.8 | 98.66 ± 6.3 |
| Metabolic Parameters, BWST |  |  |  |  |
| $\dot{V}O_{2max}$ (mL·min<sup>-1</sup>·kg<sup>-1</sup>) | **24.24 ± 7.8£** | **18.80 ± 5.4‡** | **29.67 ± 5.7‡** | **40.27 ± 7.6\*£‡** |
| RER (VCO<sub>2</sub>/VO<sub>2</sub>) | 1.04 ± 0.1 | 1.01 ± 0.0 | 1.07 ± 0.1 | 1.05 ± 0.1 |
| HR<sub>max</sub> (bpm) | 152 ± 30 | **132 ± 19‡** | 172 ± 26 | **177 ± 6.8‡** |
| % $\dot{V}O_{2max}$ | **18 ± 23£** | **3 ± 4‡** | **32 ± 25‡** | **83 ± 20\*£‡** |
| %HR<sub>max</sub> | 90.38 ± 16.8 | 83.89 ± 20.1 | 96.88 ± 11.1 | 102.95 ± 5.5 |
| Criterion for Achieving VO<sub>2</sub><sup>a</sup>, TBRS |  |  |  |  |
| RER (VCO<sub>2</sub>/VO<sub>2</sub>) ≥ 1.10 | 60 | 40 | 80 | 57 |
| Within 10% of HR<sub>max</sub><br>Predicted | 50 | 40 | 60 | 86 |
| $\dot{V}O_2$ Plateau (150mL·min<sup>-1</sup>) | 40 | 80 | 0 | 14 |
| Volitional Exhaustion | 100 | 100 | 100 | 100 |
| Achieved overall criteria for<br>$\dot{V}O_{2max}$ (≥ 2 of<br>above criterion) | 90 | 100 | 80 | 100 |
| Criterion for Achieving VO<sub>2</sub><sup>a</sup>, BWST |  |  |  |  |
| RER (VCO<sub>2</sub>/VO<sub>2</sub>) ≥ 1.10 | 20 | 0 | 40 | 43 |
| Within 10% of HR<sub>max</sub><br>Predicted | 50 | 20 | 80 | 100 |
| $\dot{V}O_2$ Plateau (150mL·min<sup>-1</sup>) | 30 | 40 | 20 | 57 |
| Volitional Exhaustion | 50 | 40 | 60 | 100 |
| Achieved overall criteria for |  |  |  |  |
| $\dot{V}O_{2max}$ ($\geq 2$ of | 60 | 40 | 80 | 100 |
| above criterion) |  |  |  |  |

Group Differences for MS Participants and Healthy Controls - EDSS, Expanded Disability Status Scale; MS, Multiple Sclerosis; RER, Respiratory Exchange Ratio; $HR_{max}$ , heart rate maximum (beats per minute, bpm); $VCO_2$ , volume of carbon dioxide; $VO_2$ , volume of oxygen; % $\dot{V}O_{2max}$ , % normative values for $\dot{V}O_{2max}$ ; % $HR_{max}$ , % age-predicted heart rate maximum. Results are reported as mean ± standard deviation, unless otherwise indicated. <sup>a</sup>, Criterion Results reported as percentage of participants who achieved the criterion. \*, Significant differences between TBRS and BWST ( $p < 0.05$ ). £, Significant differences between Controls and MS ( $p < 0.05$ ). ‡, Significant differences between the MS severity groups (EDSS $\leq 2$ , EDSS $> 2$ ) and Control

#### HR_max_and age-predicted HR_max_

In terms of HR_max_ values, there were no differences between MS and Controls (Group, F _(1,15)_ = 3.46, *p* = 0.08, µ^2^ = 0.18), between modalities (F _(1,15)_ = 0.57, *p* = 0.4, µ^2^ = 0.03), or interaction effect (F _(1,15)_ = 1.64, *p* = 0.2, µ^2^ = 0.09). The mean values of HR_max_ were only different between Control and MS subgroup EDSS > 2 (*p* = 0.002, 95% C.I. = 16.6, 73.6) on BWST but not on TBRS (F _(2,14)_ = 3.0, *p* = 0.08, µ^2^ = 0.3) (table 2). In terms of age-predicted HR_max,_, both PwMS and Controls reached 90% or greater (table 3), ([F _(1, 15)_ = 2.23, *p* = 0.1, µ^2^ = 0.13)], with no differences between the modalities [(TBRS vs, BWST), (F _(1, 15)_ = 0.49, *p* = 0.4, µ^2^ = 0.03)], or interaction effect [(F _(1, 15)_ = 1.75, *p* = 0.2, µ^2^ = 0.1]). Also, there were no differences between the PwMS subgroups: EDSS ≤ 2, EDSS > 2, and Control on either of BWST (F _(2, 14)_ = 3.26, *p* = 0.06, µ^2^ = 0.31), or TBRS (F _(2, 14)_ = 0.63, *p* = 0.5, µ^2^ = 0.08).

#### Criteria for Achieving a Maximal Cardiorespiratory Fitness Test

Overall, all Controls achieved the criteria for a V̇O_2max_ test using both the TBRS and BWST (table 4). Notably, all Control participants reached their predicted HR_max_ using BWST, and 86% did so using the TBRS. All Controls reached volitional exhaustion using both modalities. Lastly, 57% reached a VO_2_ plateau using the BWST, whereas 14% reached a VO_2_ plateau using the TBRS. Five of the 10 PwMS were unable to continue on the BWST (due to leg weakness, pain or fatigue) which precluded the ability to determine whether VO_2_ plateau had been reached (table 4). 90% of PwMS achieved the criteria for a V̇O_2max_ test (i.e., two or more of the individual criterion) using the TBRS, whereas 60% achieved the criteria using the BWST. The RER criterion was reached by 60% of PwMS using the TBRS and 20% using the BWST. In the MS subgroup with greater disability, more CPET criteria were reached using TBRS. These 5 participants reached 13/20 criteria on TBRS and only 5/20 on BWST.

**Table 4.**
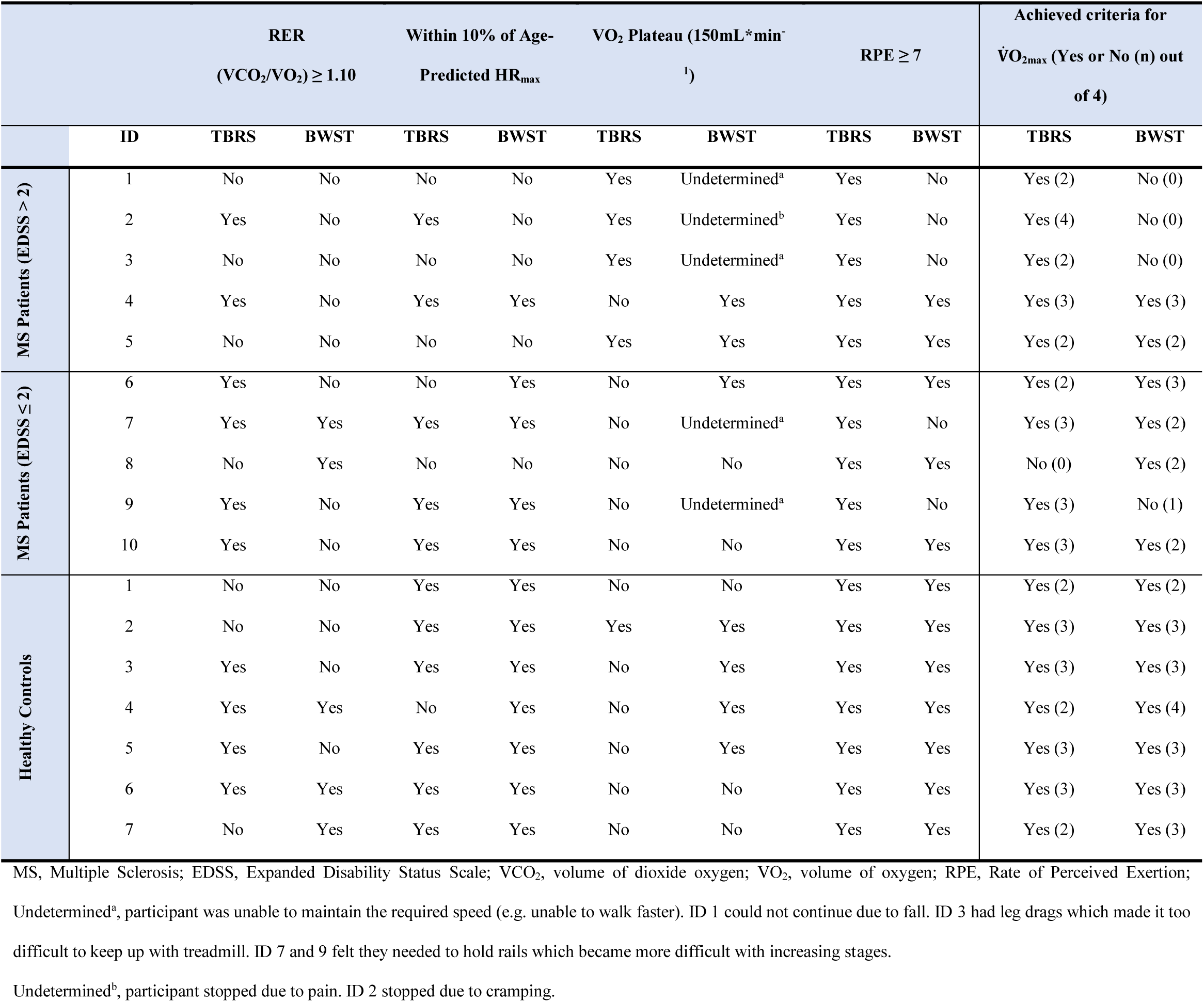
Criteria to satisfy the achievement of maximal aerobic capacity-.

## DISCUSSION

We undertook this study to optimize methods for achieving maximal CPET among PwMS with and without mobility disability, especially in longitudinal studies in which abilities, particularly walking ability, can change over time. We compared standard parameters between TBRS and BWST, including V̇O_2max_, HR_max_, age- and sex-predicted %V̇O_2max_, RER, RPE reached, age-predicted HR_max,_ and time to test completion. We also examined the criteria for reaching a maximal VO_2_ between modalities, including VO_2_ plateau (primary criterion), RER of at least 1.1, achieving at least 10% of age-predicted HR_max_, and a minimum of 7/10 RPE scale.

We report three key findings. First, contrary to our hypotheses, Controls achieved higher V̇O_2max_ and %V̇O_2max_ using BWST compared to TBRS. We also expected that PwMS would achieve higher values on TBRS but in fact, PwMS performed similarly on both devices (figures 1 and 3). This likely contributed to significantly higher V̇O_2max_ in Controls compared to PwMS only on BWST and not TBRS. Secondly, while performing BWST, CPET was compromised among PwMS since half could not continue walking due to leg symptoms, which prevented the determination of VO_2_ plateau (table 4). As for PwMS, 9/10 achieved the criteria for V̇O_2max_ (i.e., two or more of the individual criterion) using the TBRS, whereas 6/10 achieved criteria using the BWST. All Controls obtained criteria for a maximum CPET on both devices (table 4). Finally, a preliminary analysis of an MS subgroup having higher levels of mobility disability (n=5) suggested that they achieved most of the criteria for a maximum CPET using TBRS (13/20 criteria achieved (five participants * four criteria) compared to CPET using BWST (5/20 criteria achieved; table 4). Inability to meet criteria for a maximum CPET using BWST in participants having higher mobility disability likely contributed to significant differences between Controls and the PwMS having EDSS > 2 (figures 2 and 4).

Cycling ergometers and treadmills serve as gold standards for CPETs in healthy subjects [<u>33</u>], however, the recumbent seated stepper has been suggested as an ideal device for PwMS [<u>34</u>]. CPET is complicated among persons with mobility disability because lower values [<u>6</u>] could be explained by either cardiopulmonary deconditioning or limb disability [<u>35</u>]. This duality is important because the CPET is meant to measure cardiopulmonary capacity rather than leg endurance. Not all adapted CPET devices perform equally, even in apparently healthy subjects. For instance, lower V̇O_2max_ on TBRS compared to BWST, evident in our Controls, was also observed among healthy highly active participants [<u>30</u>]. Previous researchers suggested that the support provided by the TBRS seat may localize fatigue to the upper and lower limbs accounting for a 5-20% reduction in V̇O_2max_ values on TBRS [<u>36</u>]. Similarly, our results point to achievement of higher V̇O_2max_ and predicted V̇O_2max_ using BWST compared to TBRS in apparently healthy controls. Controls had little difficulty meeting criteria for CPET, the same could not be said for PwMS. They were more likely to reach criteria for a CPET on the TBRS which likely provided a more representative V̇O_2max_ value. We propose, although the PwMS performed similarly on both devices (figures 1 and 3), the TBRS more reliably represented true V̇O_2max_.

Few studies disclose whether persons with neurological disability meet predefined criteria to satisfy reaching V̇O_2max_ [<u>37</u>, <u>38</u>]. In one study, Mackay-Lyons and Makrides [<u>20</u>] reported CPET criteria performed 1-month post-stroke with 76% of patients achieving one or more of the V̇O_2max_ criteria on BWST [<u>20</u>]. Although 62% of the stroke patients achieved RER criteria on BWST [<u>20</u>], only 20% did in our patient sample. Similarly, there is a lack of information related to achieving CPET criteria in PwMS. Pilutti and group [<u>19</u>] compared TBRS and arm ergometry (EDSS=3.0), reporting that participants achieved similar V̇O_2max_ on both devices (25.2±6.8 vs. 22.5±10.1 mL*min^-1^kg^-1^) [<u>19</u>], whether participants achieved the necessary criteria for V̇O_2max_ was not reported.

The plateau in VO_2_ stands out as a primary criterion in the measurement of aerobic capacity [<u>39-41</u>], and is the best evidence that a true V̇O_2max_ is achieved [<u>42</u>]. However, VO_2_ plateau (≤150 ml/min^-1^) has been criticized because of the lack of theoretical and statistical basis, as well as its insufficient specificity to the testing protocols [<u>43</u>]. A significant variability in the percentage of subjects who showed a plateau in VO_2_ has also been reported [<u>44</u>]. In this study, achieving VO_2_ plateau was the most challenging criterion to meet using BWST or TBRS for both PwMS and Controls. Still, 30% of our PwMS achieved the VO_2_ plateau on BWST; more than the 17% reported previously in stroke [<u>20</u>] and similar to 34% of stroke patients tested on a semi-recumbent cycle ergometer [<u>45</u>]. Notably, 40% of our PwMS achieved this criteria using TBRS. Some authors propose concerns about falling may prevent PwMS from pushing themselves hard enough on the BWST [<u>16</u>]. With greater muscle fatigue on BWST [<u>21</u>], TBRS may be a better option for PwMS when conducting longitudinal studies. With challenges like the participants’ physical fitness status and the protocols used influencing the incidence of VO_2_ plateaus [<u>40</u>, <u>46</u>], further investigations need to standardize the VO_2_ plateau achievement. However, based on our results, using TBRS likely represents a true VO_2_ in PwMS (and potentially other groups experiencing mobility disability) based on VO_2_ plateau results.

Time to complete a CPET should be between 8 and 12 minutes [<u>22</u>]. Most PwMS and all but one Control finished the test within 13 and 17 minutes, respectively, using the TBRS. The time ranged from 13 to 35 minutes using BWST across both groups, suggesting that TBRS was more efficient at obtaining V̇O_2max_. Overall, older individuals or patients may require a longer time to achieve V̇O_2max_ than healthy, trained, or active subjects [<u>47</u>]. Grover et al. reported that PwMS had lower peak plantarflexor torque following 30-minute BWST exercise compared to the same intensity and duration using TBRS [<u>21</u>]. Participants in Grover’s study also took a longer time to achieve their peak torques on BWST [<u>21</u>] which may relate to leg fatigue using BWST. Leg fatigue may limit the patient’s perception of cardiopulmonary exertion on BWST [<u>12</u>, <u>16</u>] lengthening the time taken to achieve V̇O_2max_ on BWST compared to the recommended 8-12 minutes by Mezzani [<u>48</u>].

## STUDY LIMITATIONS

We provided a thorough reporting of CPET metrics in Controls and PwMS with different degrees of mobility disability. Although we provide comprehensive CPET data, the sample is small, so analyses are exploratory. Although the time to complete a CPET has been suggested to be 8-12 minutes, PwMS exercised about 9 minutes longer on BWST compared to TBRS. Therefore, fatigue may be considered as a limiting factor before cardiorespiratory exhaustion.

## CONCLUSIONS

PwMS achieved similar V̇O_2max_ and %V̇O_2max_ on the BWST and TBRS, but Controls achieved higher values using BWST. Determination of VO_2_ plateau -the main criteria- was more challenging on BWST for both groups, predictable in PwMS, probably due to MS symptoms in their legs limiting their ability to continue walking. Also, a higher percentage of PwMS achieved overall criteria for V̇O_2max_ using TBRS. All Controls achieved overall criteria on both modalities. Lastly, PwMS subgroup having higher levels of mobility disability, EDSS > 2 achieved most of the criteria for a maximum CPET using TBRS. During the preliminary investigation of PwMS subgroups with lower and higher mobility disability, CPET using BWST exaggerated already lower CPET metrics in persons with mobility disability. Based on our results, using TBRS more likely represents a true VO_2_ in PwMS (and potentially other groups experiencing mobility disability).

## Data Availability

All data produced and collected in the present study are available upon reasonable request to the authors

## LIST OF ABBREVIATIONS

ACSM: American College of Sports Medicine
BWST: Body Weight-Supported Treadmill
CPET: Cardiopulmonary Exercise Testing
EDSS: Expanded Disability Status Scale
HR_max_: the Maximum Heart Rate
MS: Multiple Sclerosis
MoCA: Montreal Cognitive Assessment
PwMS: Persons with MS
PAR-Q: Physical Activity Readiness Questionnaire
PPMS: Primary Progressive MS
RER: Respiratory Exchange Ratio
RRMS: Relapsing-Remitting MS
RPE: Rate of Perceived Exertion
SPMS: Secondary Progressive MS
TBRS: Total Body Recumbent Stepper
V̇O_2max_: the Maximum Rate of Oxygen Consumption
V̇CO_2_: Volume of Carbon Dioxide Exhaled
V̇O2: Volume of Oxygen Inhaled

## HIGHLIGHTS

- Testing aerobic fitness accurately is crucial in persons with multiple sclerosis.
- There is no evidence-based consensus regarding the best testing modality that should preferentially be used in patients with mobility disability.
- Comparing two modalities (body weight-supported treadmill vs. total body recumbent stepper) showed that patients achieve more of the criteria required to satisfy an actual maximal aerobic capacity on the stepper.
- Considering the leg symptoms in MS patients, testing aerobic capacity using a stepper is suggested.

## DECLARATION OF INTEREST

The authors declared no potential conflicts of interest related to the research, authorship, and publication.

## FUNDING

This research was supported by MS Canada, Grant#2631 (MP); Canada Research Chairs Program Grant #950-232532 (MP) and Canada Foundation for Innovation Grant #211716 (MP).

## ACKNOWLEDGMENTS

The authors acknowledge the participants for donating their time.

## Notes

### Competing Interest Statement

The authors have declared no competing interest.

### Funding Statement

This research was supported by MS Canada, Grant#2631; Canada Research Chairs Program Grant #950-232532; and Canada Foundation for Innovation Grant #211716.

### Author Declarations

The Health Research Ethics Board (HREB) of Memorial University of Newfoundland gave ethical approval for this study (Approval Ref. #: 14.102).

## REFERENCES

1. Koch-Henriksen, N. and Sorensen, P.S. The changing demographic pattern of multiple sclerosis epidemiology. Lancet Neurol, 2010. 9(5): p. 520–32. 10.1016/S1474-4422(10)70064-8.

2. Ann Marrie, R. and R.I. Horwitz, Emerging effects of comorbidities on multiple sclerosis. Lancet Neurology, 2010. 9(8): p. 820–828. 10.1016/S1474-4422(10)70135-6.

3. Wens, I. Dalgas, U. Stenager, E. Eijnde, B. O. Risk factors related to cardiovascular diseases and the metabolic syndrome in multiple sclerosis - a systematic review. Multiple Sclerosis Journal, 2013. 19(12): p. 1556–1564. DOI: 10.1177/1352458513504252.

4. Eriksen, L. Curtis, T. Grønbæk, M. Helge, J. W. Tolstrup, J. S. The association between physical activity, cardiorespiratory fitness and self-rated health. Preventive Medicine, 2013. 57(6): p. 900–2. DOI: 10.1016/j.ypmed.2013.09.024.

5. Bassett JR, D.R. and E.T. Howley, Limiting factors for maximum oxygen uptake and determinants of endurance performance. Medicine and Science in Sports and Exercise, 2000. 32(1): p. 70–84.

6. Langeskov-Christensen, M. Heine, M. Kwakkel, G. Dalgas, U. Aerobic capacity in persons with multiple sclerosis: a systematic review and meta-analysis. Sports Medicine, 2015. 45(6): p. 905–23. DOI: 10.1007/s40279-015-0307-x.

7. Guerra, E.d.C., A. Mancini, P. Sperandii, F. Quaranta, F. Ciminelli, E. Fagnani, F. Giombini, A. Pigozzi, F., Physical fitness assessment in multiple sclerosis patients: a controlled study. Research in Developmental Disabilities, 2014. 35(10): p. 2527–33. DOI: 10.1016/j.ridd.2014.06.013.

8. Sandroff, B.M., J.J. Sosnoff, and R.W. Motl, Physical fitness, walking performance, and gait in multiple sclerosis. Journal of the Neurological Sciences, 2013. 328(1-2): p. 70–76. DOI: 10.1016/j.jns.2013.02.021.

9. Sandroff, B.M. and R.W. Motl, Fitness and Cognitive Processing Speed in Persons with Multiple Sclerosis: A Cross-Sectional Investigation. Journal of Clinical and Experimental Neuropsychology 2012. 34(10): p. 1041–1052. DOI: 10.1080/13803395.2012.715144.

10. Prakash, R.S. Snook, E. M. Motl, R. W. Kramer, A. F. Aerobic fitness is associated with gray matter volume and white matter integrity in multiple sclerosis. Brain Research, 2010. 1341: p. 41–51. DOI: 10.1016/j.brainres.2009.06.063.

11. van den Akker, L.E. Heine, M. van der Veldt, N. Dekker, J. de Groot, V. Beckerman, H. Feasibility and Safety of Cardiopulmonary Exercise Testing in Multiple Sclerosis: A Systematic Review. Archives of Physical Medicine and Rehabilitation, 2015. 96(11): p. 2055–2066. DOI: 10.1016/j.apmr.2015.04.021.

12. Pilutti, L.A. Paulseth, J. E. Dove, C. Jiang, S. Rathbone, M. P. Hicks, A. L. Exercise Training in Progressive Multiple Sclerosis: A Comparison of Recumbent Stepping and Body Weight-Supported Treadmill Training. International Journal of MS Care, 2016. 18(5): p. 221–229. DOI: 10.7224/1537-2073.2015-067.

13. Valet, M. Lejeune, T. Hakizimana, J. C. Stoquart, G. Quality of the tools used to assess aerobic capacity in people with multiple sclerosis. European Journal of Physical and Rehabilitation Medicine, 2017. 53(5): p. 759–774. DOI: 10.23736/S1973-9087.17.04218-6.

14. Rampello, A. Franceschini, M. Piepoli, M. Antenucci, R. Lenti, G. Olivieri, D. Chetta, A. Effect of Aerobic Training on Walking Capacity and Maximal Exercise Tolerance in Patients With Multiple Sclerosis: A Randomized Crossover Controlled Study. American Physical Therapy Association, 2007. 87: p. 545–555. 10.2522/ptj.20060085.

15. Dawes, H. Collett, J. Meaney, A. Duda, J. Sackley, C. Wade, D. Barker, K. Izadi, H. Delayed recovery of leg fatigue symptoms following a maximal exercise session in people with multiple sclerosis. Neurorehabilitation and Neural Repair, 2014. 28(2): p. 139–48. DOI: 10.1177/1545968313503218.

16. Kalron, A. and A. Achiron, The relationship between fear of falling to spatiotemporal gait parameters measured by an instrumented treadmill in people with multiple sclerosis. Gait Posture, 2014. 39(2): p. 739–44. DOI: 10.1016/j.gaitpost.2013.10.012.

17. Salisbury, D.L. Swanson, K. Brown, R. J. Treat-Jacobson, D. Total body recumbent stepping vs treadmill walking in supervised exercise therapy: A pilot study. Vascular Medicine, 2022. 27(2): p. 150–157. DOI: 10.1177/1358863X211068888.

18. Devasahayam, A.J. Chaves, A. R. Lasisi, W. O. Curtis, M. E. Wadden, K. P. Kelly, L. P. Pretty, R. Chen, A. Wallack, E. M. Newell, C. J. Williams, J. B. Kenny, H. Downer, M. B. McCarthy, J. Moore, C. S. Ploughman, M. Vigorous cool room treadmill training to improve walking ability in people with multiple sclerosis who use ambulatory assistive devices: a feasibility study. BMC Neurol, 2020. 20(1): p. 33. DOI: 10.1186/s12883-020-1611-0.

19. Pilutti, L.A. Sandroff, B. M. Klaren, R. E. Learmonth, Y. C. Platta, M. E. Hubbard, E. A. Stratton, M. Motl, R. W. Physical Fitness Assessment Across the Disability Spectrum in Persons With Multiple Sclerosis: A Comparison of Testing Modalities. Journal of Neurologic Physical Therapy, 2015. 39(4): p. 241–249. DOI: 10.1097/NPT.0000000000000099.

20. Mackay-Lyons, M.J. and L. Makrides, Exercise capacity early after stroke. Archives of Physical Medicine and Rehabilitation, 2002. 83(12): p. 1697–702. DOI: 10.1053/apmr.2002.36395.

21. Grover, G. Ploughman, M. Philpott, D. T. Kelly, L. P. Devasahayam, A. J. Wadden, K. Power, K. E. Button, D. C. Environmental temperature and exercise modality independently impact central and muscle fatigue among people with multiple sclerosis. Multiple Sclerosis Journal, 2017. 3: p. 4. DOI: 10.1177/2055217317747625.

22. Liguori G. and Wolters K. American College of Sports Medicine Guidelines for Exercise Testing and Prescription. 11 ed, ed. 2022.

23. Ponichtera-Mulcare, J.A. Mathews, T. Glaser, R. M. Gupta, S. C. Maximal Aerobic Exercise of Individuals with Multiple Sclerosis Using Three Modes of Ergometry. Clinical Kinesiology 1995. 49(1): p. 4–13.

24. White, A.T. Wilson, T. E. Davis, S. L. Petajan, J. H. Effect of precooling on physical performance in multiple sclerosis. Multiple Sclerosis Journal, 2000. 6(3): p. 176–180. DOI: 10.1177/135245850000600307.

25. Polman, C.H. Reingold, S. C. Banwell, B. Clanet, M. Cohen, J. A. Filippi, M. Fujihara, K. Havrdova, E. Hutchinson, M. Kappos, L. Lublin, F. D. Montalban, X. O’Connor, P. Sandberg-Wollheim, M. Thompson, A. J. Waubant, E. Weinshenker, B. Wolinsky, J. S. Diagnostic criteria for multiple sclerosis: 2010 revisions to the McDonald criteria. Annals of Neurology, 2011. 69(2): p. 292–302. DOI: 10.1002/ana.22366.

26. Bredin, S.S.D. Gledhill, N. Jamnik, V. K. Warburton, D.E.R. PAR-Q+ and ePARmed-X+ New risk stratification and physical activity clearance strategy for physicians and patients alike. Canadian Family Physician, 2013. 59(3): p. 273–277.

27. Nasreddine, Z.S., et al., The Montreal Cognitive Assessment, MoCA: a brief screening tool for mild cognitive impairment. Journal of the American Geriatrics Society, 2005. 53(4): p. 695–699. DOI: 10.1111/j.1532-5415.2005.53221.x.

28. Crystal, O., Kean, D.G. Behm, and W.B. Young, Fixed Foot Balance Training Increases Rectus Femoris Activation During Landing And Jump Height in Recreationally Active Women. Journal of Sports Science and Medicine, 2006. 5: p. 138–148.

29. Chaves, A.R., et al., Exercise-Induced Brain Excitability Changes in Progressive Multiple Sclerosis: A Pilot Study. Journal of Neurologic Physical Therapy, 2020. 44(2): p. 132–144. DOI: 10.1097/NPT.0000000000000308

30. Billinger, S.A., B.Y. Tseng, and P.M. Kluding, Modified Total-Body Recumbent Stepper Exercise Test for Assessing Peak Oxygen Consumption in People With Chronic Stroke. Physical Therapy, 2008. 88: p. 7. DOI: 10.2522/ptj.20080072.

31. Ferguson, B., ACSM’s Guidelines for Exercise Testing and Prescription 9th Ed. 2014. The Journal of the Canadian Chiropractic Association, 2014. 58(3): p. 328.

32. Bakeman, R., Recommended effect size statistics for repeated measures designs. Behavior Research Methods, 2005. 37(3): p. 379–384. DOI: 10.3758/BF03192707.

33. Motl, R.W. and B. Fernhall, Accurate prediction of cardiorespiratory fitness using cycle ergometry in minimally disabled persons with relapsing-remitting multiple sclerosis. Archives of Physical Medicine and Rehabilitation, 2012. 93(3): p. 490–495. DOI: 10.1016/j.apmr.2011.08.025.

34. Motl, R.W., et al., Cardiorespiratory fitness and its association with thalamic, hippocampal, and basal ganglia volumes in multiple sclerosis. Neuroimage Clinical, 2015. 7: p. 661–666.DOI: 10.1016/j.nicl.2015.02.017.

35. Motl, R.W. and M. Goldman, Physical inactivity, neurological disability, and cardiorespiratory fitness in multiple sclerosis. Acta Neurologica Scandinavica, 2011. 123(2): p. 98–104. DOI: 10.1111/j.1600-0404.2010.01361.x.

36. Gordon, N.F., et al., Physical activity and exercise recommendations for stroke survivors: an American Heart Association scientific statement from the Council on Clinical Cardiology, Subcommittee on Exercise, Cardiac Rehabilitation, and Prevention; the Council on Cardiovascular Nursing; the Council on Nutrition, Physical Activity, and Metabolism; and the Stroke Council. Circulation, 2004. 109(16): p. 2031–41. DOI: 10.1161/01.CIR.0000126280.65777.A4.

37. van de Port, I.G., G. Kwakkel, and H. Wittink, Systematic review of cardiopulmonary exercise testing post stroke: Are we adhering to practice recommendations? Journal of Rehabilitation Medicine, 2015. 47(10): p. 881–900. DOI: 10.2340/16501977-2031.

38. Ploughman, M., et al., Synergistic Benefits of Combined Aerobic and Cognitive Training on Fluid Intelligence and the Role of IGF-1 in Chronic Stroke. Neurorehabil Neural Repair, 2019. 33(3): p. 199–212. DOI: 10.1177/1545968319832605.

39. Astorino, T.A., Alterations in VO2max and the VO2 plateau with manipulation of sampling interval. Clinical Physiology and Functional Imaging, 2009. 29: p. 60–67. DOI: 10.1055/s-0028-1104588.

40. Midgley AW and C. S., Emergence of the verification phase procedure for confirming ‘true’ VO(2max). Scandinavian Journal of Medicine & Science in Sports, 2009. 19(3): p. 313–322.

41. Poole, D.C. and A.M. Jones, Measurement of the maximum oxygen uptake V̇o2max: V̇o2peak is no longer acceptable. Journal of Applied Physiology, 2017. 122(4): p. 997–1002. DOI: 10.1152/japplphysiol.01063.2016.

42. Albouaini K, et al., Cardiopulmonary exercise testing and its application. Heart, 2007. 93(10): p. 1285–1292. DOI: 10.1136/hrt.2007.121558.

43. Astorino TA, et al., Incidence of the oxygen plateau at VO2max during exercise testing to volitional fatigue. Journal of Exercise Physiology Online, 2000. 3(4): p. 12.

44. Mier, C.M., R.P. Alexander, and A.L. Mageean, Achievement of VO2max criteria during a continuous graded exercise test and a verification stage performed by college athletes. The Journal of Strength & Conditioning Research, 2012. 26(10): p. 2648–2654. DOI: 10.1519/JSC.0b013e31823f8de9.

45. Tang, A., et al., Maximal exercise test results in subacute stroke. Arch Phys Med Rehabil, 2006. 87(8): p. 1100–5. DOI: 10.1016/j.apmr.2006.04.016.

46. Astorino, T.A., A.C. White, and L.C. Dalleck, Supramaximal testing to confirm attainment of VO2max in sedentary men and women. International Journal of Sports Medicine, 2009. 30(4): p. 279–84. DOI: 10.1111/j.1475-097X.2008.00835.x.

47. Schaun, G.Z., The Maximal Oxygen Uptake Verification Phase: a Light at the End of the Tunnel? Sports Medicine-Open, 2017. 3(1): p. 44. DOI: 10.1186/s40798-017-0112-1.

48. Mezzani, A., Cardiopulmonary Exercise Testing: Basics of Methodology and Measurements. Annals of the American Thoracic Society, 2017. 14(Supplement_1): p. S3-S11. DOI: 10.1513/AnnalsATS.201612-997FR.

